# Is higher-level trauma center care associated with better outcomes in patients injured by low-energy trauma?

**DOI:** 10.1101/2021.06.01.21258178

**Authors:** Michael Tonkins, Daniel Bradbury, Paul Bramley, Lisa Sabir, Anna Wilkinson, Fiona Lecky

**Affiliations:** School of Health and Related Research, University of Sheffield, Sheffield, UK; Sheffield Children’s Hospital NHS Foundation Trust, Sheffield, UK; Sheffield Teaching Hospitals NHS Foundation Trust, Sheffield, UK

**Keywords:** Trauma, trauma systems, falls

## Abstract

**Background:** In high-income countries trauma patients are becoming older, more likely to have comorbidities, and are being injured by low-energy mechanisms, chiefly ground-level falls. It is currently unknown whether existing trauma systems improve the outcomes of these patients. This systematic review investigates the association between higher-level trauma center care and outcomes of adult patients who were admitted to hospital due to injuries sustained following low-energy trauma.

**Methods:** A pre-registered systematic review (CRD42020211652) of subject databases and grey literature, supplemented by targeted manual searching, was conducted in January 2021. Studies were eligible if they reported outcomes in adults admitted to hospital due to low-energy trauma. Studies were excluded if participants were not adults or were not admitted to hospital. Studies in lower- and middle-income settings were excluded due to differences in demographics and healthcare systems. Risk of bias was assessed by independent reviewers using the Robins-I tool. In the presence of study heterogeneity a narrative synthesis was pre-specified.

**Results:** Three observational studies were included from 2,898 unique records. The studies’ risk of bias was moderate-to-serious due to potential residual confounding and selection bias. All studies compared outcomes among adults injured by ground-level falls treated in trauma centers verified by the American College of Surgeons in the USA. The studies reported divergent results. One demonstrated improved outcomes in level 3 or 4 trauma centers (Observed: Expected Mortality 0.973, 95%CI 0.971-0.975), one demonstrated improved outcomes in level 1 trauma centers (Adjusted Odds Ratio 0.71, 95%CI 0.56-0.91), and one demonstrated no difference between level 1 or 2 and level 3 or 4 trauma center care (Adjusted Odds Ratio 0.91 (0.80-1.04).

**Conclusions:** There is currently no strong evidence for the efficacy of major trauma centers in caring for adult patients injured by a ground-level fall. Further studies at lower risk of bias and studies conducted outwith the USA are required.

**Level of Evidence:** Level III systematic review and meta-analysis

## Introduction

Trauma has previously been considered a disease of the young. Epidemiological studies have emphasized the loss of life and prevalence of disability in working-age people due to road traffic injuries, self-inflicted injuries and interpersonal violence.^1,2^ Consequently, trauma systems have been developed in many countries to afford rapid access to resuscitation facilities and specialized care.^3^

In general, trauma systems consist of a higher-level center with full facilities and specialist services; nearby lower-level centers with less comprehensive facilities; and pre-hospital services which transport injured patients to hospital based on field triage criteria. A description of trauma center levels in the USA is given in Box 1. Although the structure and function of trauma systems is heterogenous there is good evidence that they are associated with lower rates of patient mortality. ^4–8^ It is essential that trauma systems are continuously monitored, and timely, relevant information about their performance returned to the different groups that care for injured patients.^9^

However, in high income countries the epidemiology and etiology of trauma is changing. The mean age of trauma patients is increasing, and so is the prevalence of comorbidities and frailty. Furthermore, high-energy mechanisms of injury such as road traffic collisions are increasingly becoming displaced by low-energy mechanisms, chiefly ground-level falls.^10^

It is well-recognized that this emerging population of trauma patients is vulnerable, with increasing age and comorbidity associated with greater risk of mortality. There is evidence that existing trauma systems may not be ideally suited to care for this changing patient group. In the prehospital environment there are high rates of undertriage,^11^ and older patients are less likely to receive major trauma center care at any point in their care journey.^12,13^ Current injury severity scores may under-estimate patient injury burden,^14^ and significant residual disability may impair the quality of life of survivors.^15^

Conversely, there is also evidence that specific hospital-based interventions or models of care can improve outcomes in older trauma patients.^16,17^

A systematic review of the performance of prehospital triage tools and risk factors in elderly patients is already underway.^18^ Therefore, the present review will address the hospital component of trauma systems by evaluating whether higher-level trauma center care is associated with improved outcomes compared to lower-level trauma center care.

**Level 1:** capable of providing total care for every aspect of injury, from prevention to rehabilitation. Level 1 centres are responsible for providing leadership in education, research, and system planning.

**Level 2:** capable of providing initial definitive care except for particularly complex or specialised injuries.

**Level 3:** capable of providing prompt assessment, resuscitation, stabilisation and transfer of patients when required.

**Level 4:** provides advanced trauma life support in remote areas where no higher level of care is available.

Box 1: Definitions of Trauma Centers Verified by the American College of Surgeons.^19^

In the United Kingdom, ‘major trauma centers’ would be equivalent to a Level 1 or 2 trauma centre, and ‘trauma units’ equivalent to a Level 3 trauma center.

### Review Question

In adults requiring hospital admission for the management of injuries sustained through low-energy trauma, is higher-level trauma center care associated with improved patient outcomes compared to care in a lower-level center?

## Methods

### Review Management

A detailed review protocol was written and registered with PROSPERO (CRD42020211652). The advice of an information specialist was sought during development of the search strategy.

### Information Sources and Search

Keyword and database heading searches were performed on terms related to trauma centers, low-energy trauma and falls. The searches were re-run immediately prior to the final analysis run on 12 January 20201. The complete search strategy is available in Supplemental Table 1.

The databases searched were MEDLINE, EMBASE and CINHAL, the Cochrane Database of Systematic Reviews and the Cochrane Central Register of Controlled Trials. Searches for grey literature were conducted on the websites OpenGrey and the Grey Literature Report.

The reference lists of included studies were hand-searched to identify antecedent studies. Online citation data was reviewed to identify potentially relevant subsequent studies.

Searches were limited from the year 1987 to present because the American College of Surgeons (ACS) Verification, Review and Consultation (VRC) Program, which established a benchmark for different ‘levels’ of trauma center, commenced in 1987.^19^ In addition, searches were limited to English language articles due to the lack of facility for translation, and to studies in humans.

### Eligibility Criteria

The population of interest was adult patients (as defined by the included studies) who were admitted to hospital due to low-energy trauma. Low-energy trauma was defined as a fall from less than 2 meters. Important synonyms included ‘low fall’, ‘ground-level fall’ and ‘fall from standing height’.

Patients were excluded if they were not adults or were not admitted to hospital. Studies in lower and middle-income settings were excluded due to the differences in demographics, injury patterns and healthcare systems.

The intervention under investigation was higher-level trauma center care. Variation in national systems precludes precise definition of exactly what this care comprises. For the purposes of this review, studies’ description of a facility as a ‘major trauma center’ was accepted, with the explicit assumption that such facilities would be on par with ACS-certified level 1 or level 2 trauma centers, or with Major Trauma Centers in the UK.^20,21^

The comparison was care in a lower-level trauma center. Similar difficulties regarding precise definition of a non-MTC apply. Once more, studies’ description of the facility was taken at face value, with the assumption that such facilities would be on par with ACS-certified level 3 and level 4 trauma centers, or Trauma Units in the UK.

Published and unpublished randomized controlled trials and non-randomized studies were eligible for inclusion. Studies were excluded at full text review if they were at high risk of bias or severely methodologically flawed.

### Outcomes

The primary outcome of interest was mortality, at the time point specified by the included study.

Secondary outcomes included morbidity, length of hospital stay, length of critical care stay, in-hospital complications and surgical operation rates.

### Selection of Studies

All identified records were pooled and duplicates removed. A single reviewer (MT) screened all titles and abstracts to establish potential eligibility. Full texts of potentially eligible studies were reviewed by two independent reviewers. Discrepancies were resolved through discussion and the adjudication of the senior author (FL).

### Quality Assessment

All eligible studies were evaluated for methodological quality and overlap of the study populations. If this was found to be the case, or if serious methodological flaws or reporting inconsistencies were identified, then studies were excluded. Risk of bias assessment was conducted by two independent reviewers using the Robins-I tool.^22^ Any discrepancies were resolved through consensus.

### Data Extraction, Synthesis and Analysis

Study-level data were extracted into a standardized data extraction form (Excel, Microsoft, Redmond) and checked by an independent reviewer. A summary of the variables extracted is in Table 1. If data fields were not presented, but could be calculated, then this was performed in Excel. All corresponding authors were contacted for additional information or clarification.

**Table 1:** Summary of data extracted from included studies.

| Table 1: Summary of data extracted from included studies |  |  |  |
| --- | --- | --- | --- |
| Study information | Population data | Process data | Outcome data |
| Author | Sample size | Intubation rate | Mortality |
| Year of publication | Age | Critical care admission | Morbidity |
| Funding source | Gender |  | Length of stay |
| Setting | Mechanism of injury |  | Critical care length of stay |
| Study design | Vital signs |  | Effect measures |
| Inclusion criteria | Injury distribution |  |  |
| Exclusion criteria | Trauma severity score |  |  |
| Recruitment method | Comorbidities |  |  |
| Definition of low energy | Anticoagulation |  |  |
|  | Alcohol or drug use |  |  |

### Patient and Public Involvement

In 2017 the Emergency Medicine Priority Setting Partnership, involving patients and careers across the UK, established that identifying which patients should receive immediate major trauma center care is the tenth most important emergency medicine research priority.^23^

## Results

### Study Selection

Searches identified 2,851 database records and 30 records from grey literature archives. After removing duplicates 2,144 unique records were screened for eligibility. Of these, 48 full papers were retrieved and 6 were found to be appropriate to proceed for quality assessment. Secondary searches of these six papers identified a further 17 potentially relevant studies. These full papers were retrieved however none were eligible. Figure 1 illustrates the process of study selection in detail.

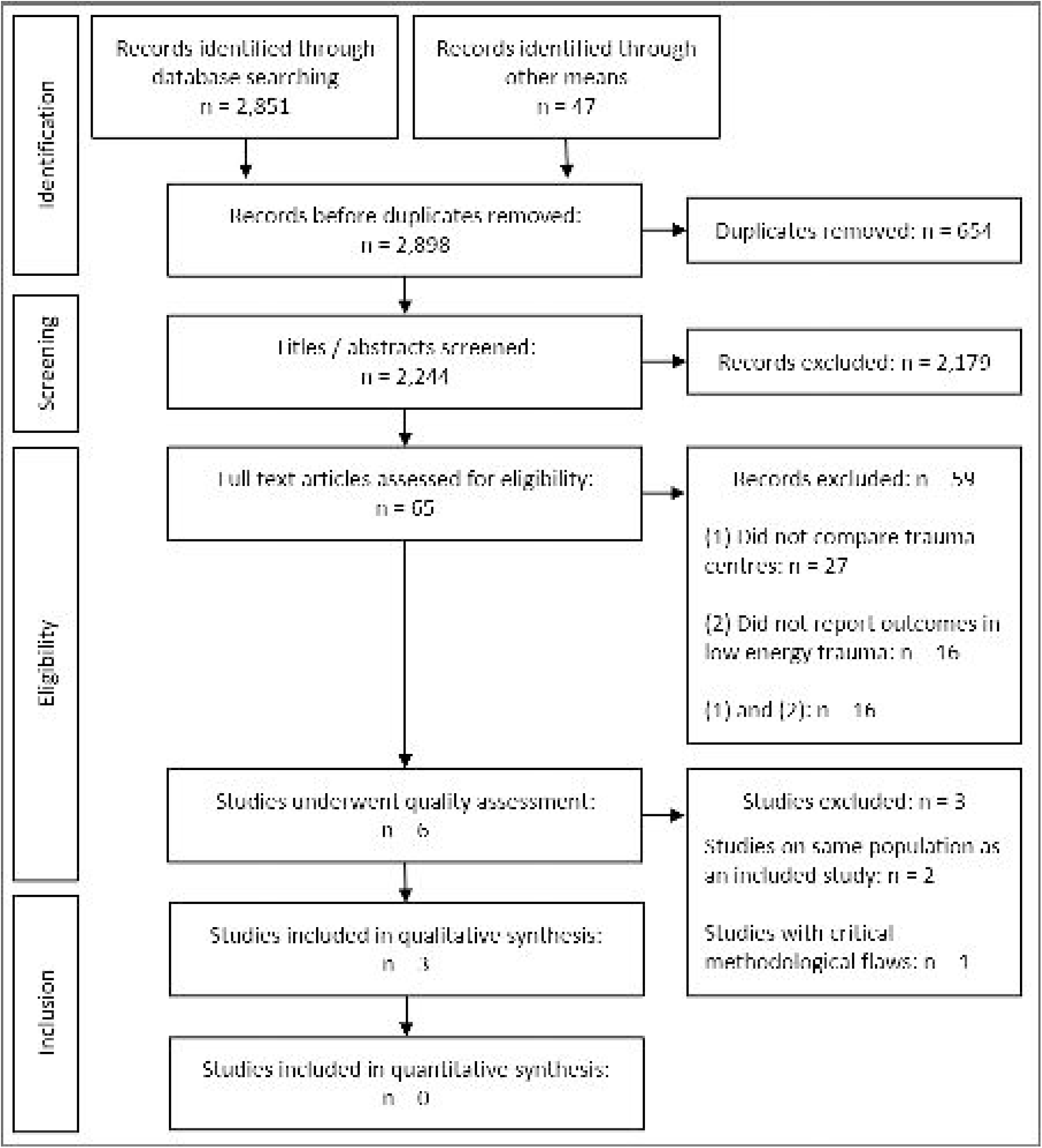

### Global Quality Assessment

Global quality assessment identified three studies that were not suitable to proceed to risk of bias assessment. One study was excluded at this point because it was found to have critical methodological flaws.^24^ Two further studies were excluded because they repeated findings in the same patient cohort,^25^ or a subset of the same cohort,^26^ as a third (included) study.^27^ Full details are given in Supplemental Table 2.

### Study Characteristics

The three included studies are summarized in Table 2.^27–29^ All were cohort studies conducted in the United States between 2001 and 2014. Two used the National Trauma Data Bank, and one used a state trauma registry. All studies compared ACS-verified level 1 and/or level 2 trauma centers (‘higher-level care’) to ACS-verified level 3 and 4 centers (‘lower-level care’).

**Table 2:** Characteristics of included studies.

| Table 2: Characteristics of included studies |  |  |  |  |  |  |  |  |  |  |
| --- | --- | --- | --- | --- | --- | --- | --- | --- | --- | --- |
| Author<br>Date | Design<br>Period | Setting<br>Data | Inclusion<br>Criteria | Exclusion<br>Criteria | Comparisons | Patients |  | Trauma Centres |  | Outcomes |
|  |  |  |  |  |  | n | % | n | % |  |
| Ahmed<br>2019<br><sup>27</sup> | RCS<br>2012 -<br>2014 | USA<br>NTDB | Age >65<br>Ground level fall<br>Fell in private residence<br>SPB 90-160mmHg<br>HR 60-100bpm<br>GCS 15 | Age <65<br>Fall "from a great height"<br>Any other mechanism<br>Patients not admitted to<br>hospital | Whole study | 40,800 | 100 | NR* |  | In-hospital mortality<br>Time to expiration<br>Total length of stay<br>Patient discharge<br>disposition |
|  |  |  |  |  | ACS 1 & 2 | 19,290 | 47.3 | NR* |  |  |
|  |  |  |  |  | ACS 3 & 4 &<br>unranked | 21,510 | 52.7 | NR* |  |  |
| Cook<br>2012<br><sup>28</sup> | RCS<br>2001 -<br>2009 | Texas<br>TSA-G | Age >14 years<br>Ground level fall<br>Admitted to hospital | Age <14 years<br>Not admitted to hospital | Whole study | 8,202 | 100 | 20 | 100 | In-hospital mortality |
|  |  |  |  |  | ACS 1 | 803 | 9.8 | 1 | 5.0 |  |
|  |  |  |  |  | ACS 2 | 2,205 | 26.9 | 2 | 10.0 |  |
|  |  |  |  |  | ACS 3 & 4 | 5,194 | 63.3 | 17 | 85.0 |  |
| Roubik<br>2017<br><sup>29</sup> | RCS<br>2007 -<br>2014 | USA<br>NTDB | Age >15 years<br>Ground level fall | Age <15 years<br>Any other mechanism<br>Missing values (age,<br>TMPM, trauma centre<br>level, comorbidities)<br>Trauma centre in lowest<br>decile of ground level falls<br>Admission <24 hours | Whole study | 554,686 | 100 | 446 | 100 | In-hospital mortality |
|  |  |  |  |  | ACS 1 & 2 | 502,587 | 90.6 | 335 | 75.1 |  |
|  |  |  |  |  | ACS 3 & 4 | 52,099 | 9.4 | 111 | 24.9 |  |
| RCS: retrospective cohort study, SBP: systolic blood pressure, NTDB: National Trauma Data Bank (34), TMPM: Trauma Mortality Prediction Model, GCS: Glasgow Coma Score, TSA-G: Regional Trauma Registry for Trauma Service Area G (Northeastern Texas). *Author contacted but data not available |  |  |  |  |  |  |  |  |  |  |

In addition, Roubik 2019 compared outcomes in state-designated trauma centers. The authors decided to separate state-designated from ACS-verified centers to “avoid heterogeneity introduced by variable criteria among states’ designation requirements” (p 37). This review reports only the outcomes in the ACS-verified centers to maintain comparability between studies. Full results are reported in Supplemental Table 3.

Whilst Cook 2012 and Roubik 2017 include all adult patients (defined as age >14 years and age >15 years respectively), Ahmed 2019’s analysis is limited to older patients (age >65 years) with normal vital signs at the scene.

There are two periods of overlapping dates covered by the studies: 2007-2009 and 2012-2014. In the first period it is likely that a proportion of the patients in the study by Cook 2012 will be included in the NTDB patients analyzed by Roubik 2017. In the second period all the patients analyzed by Ahmed 2019 would also be expected to be analyzed by Roubik 2017.

All studies used inpatient mortality as the primary outcome, and only Ahmed 2019 reported secondary outcomes.

### Risk of Bias

Risk of bias assessment is summarized in Table 3.

**Table 3:**
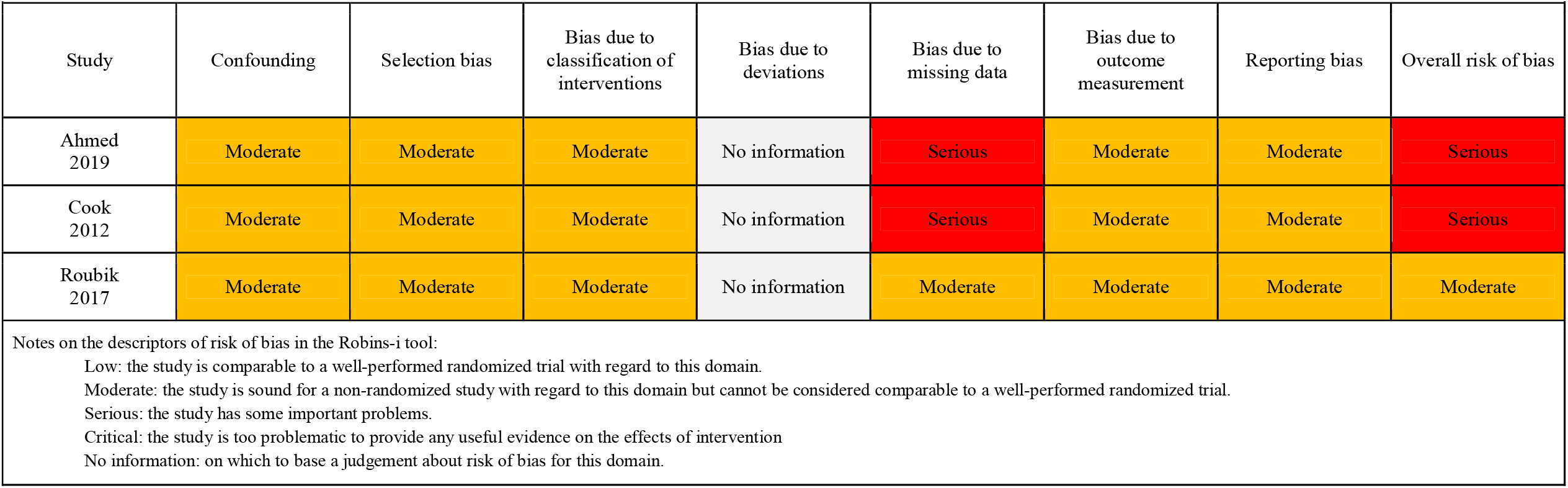
Risk of bias assessment using the Robins-i tool.

| Study | Confounding | Selection bias | Bias due to classification of interventions | Bias due to deviations | Bias due to missing data | Bias due to outcome measurement | Reporting bias | Overall risk of bias |
| --- | --- | --- | --- | --- | --- | --- | --- | --- |
| Ahmed 2019 | Moderate | Moderate | Moderate | No information | Serious | Moderate | Moderate | Serious |
| Cook 2012 | Moderate | Moderate | Moderate | No information | Serious | Moderate | Moderate | Serious |
| Roubik 2017 | Moderate | Moderate | Moderate | No information | Moderate | Moderate | Moderate | Moderate |

Notes on the descriptors of risk of bias in the Robins-i tool:
Low: the study is comparable to a well-performed randomized trial with regard to this domain.
Moderate: the study is sound for a non-randomized study with regard to this domain but cannot be considered comparable to a well-performed randomized trial.
Serious: the study has some important problems.
Critical: the study is too problematic to provide any useful evidence on the effects of intervention
No information: on which to base a judgement about risk of bias for this domain.

Selection of participants into each study was judged to be at moderate risk of bias. This was largely due to the potential for confounding by indication, which may have occurred in the pre-hospital setting, or tertiary referral bias, which may have arisen due to the transfer of patients from lower-level centers to higher-level centers. The handling of patient transfers between centers is also of particular concern because this has been found to be associated with better patient outcomes.^30,31^ Ahmed 2019 does not describe how transferred patients were analyzed. In both Cook 2012 and Roubik 2017 transferred patients were assigned to the receiving trauma center.

All studies used appropriate methods to attempt to control for confounding factors using multiple logistic regression (Cook 2012, Roubik 2017) or nearest neighbor matching (Ahmed 2019). The models are compared in Supplemental Table 4. Briefly, the models comprise a mixture of demographic data, clinical observations, patient comorbidities, injury distribution, trauma severity scores (trauma mortality prediction index or injury severity score) and the results of investigations. Ahmed 2019 used 7 variables, Cook 2012 used 13 variables and Roubik 2017 used 46 variables.

In general, the reliance upon trauma databanks may introduce bias due to unmeasured, missing or inaccurate data.^32^ Two studies were judged as being at ‘serious’ risk of bias due to the handling of missing data. Ahmed 2019 acknowledges that missing data could influence their propensity score matching, but provide no account of the nature, quantity or distribution of missingness. Cook 2012 makes no reference to missing data. Roubik 2017 found 1% of patients were missing data for predictor variables and tested the implications of this, finding that it made no significant difference to their results. However the data were not missing at random (level 3 and 4 centers had a higher rate of missing data) and therefore the study was conservatively judged to be at moderate risk of bias.

Finally, all studies were at risk of reporting bias due to the lack of published pre-specified analysis plans.

### Mortality by Trauma Centre Level

Cook 2012 and Roubik 2017 both report unadjusted mortality by trauma center level, demonstrating higher unadjusted mortality at higher-level trauma centers (Table 4). After adjustment the findings for the primary outcome of each study are divergent.

**Table 4:**
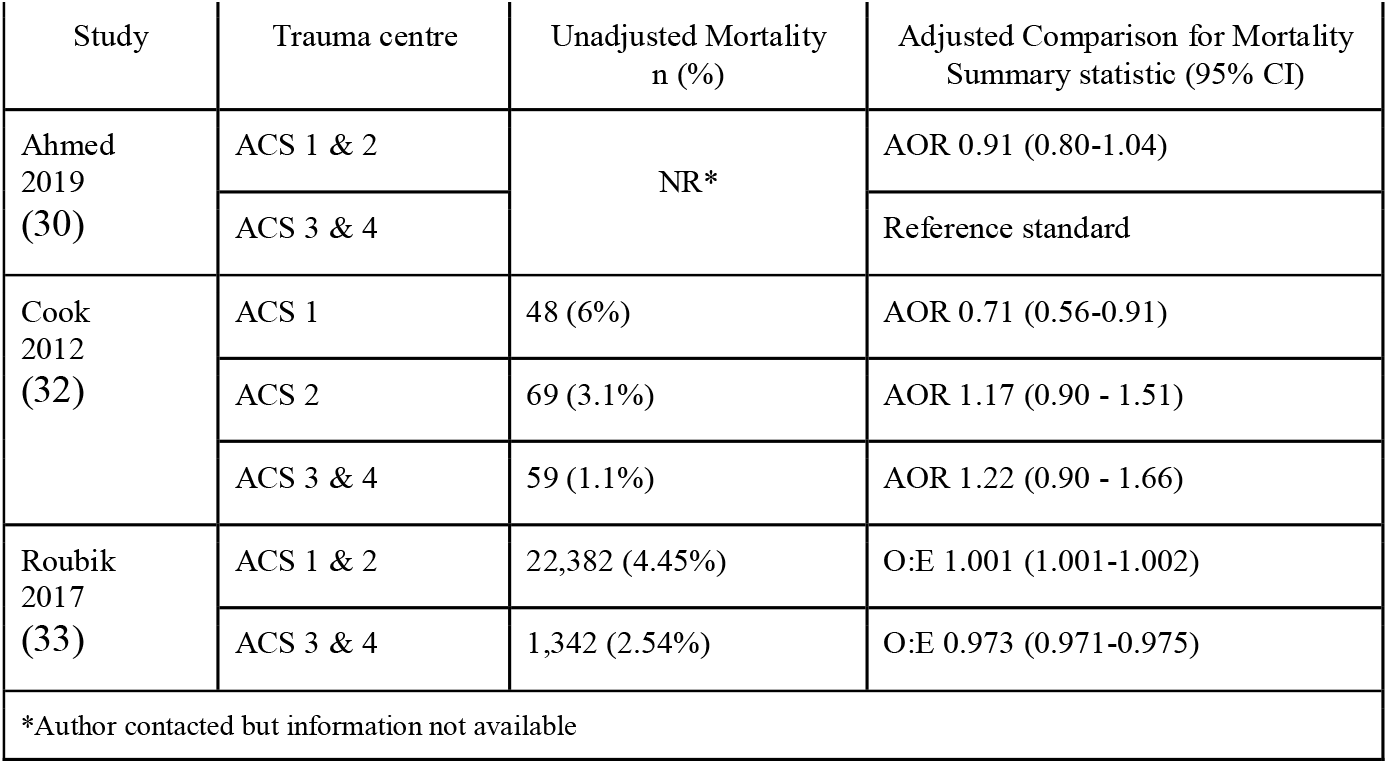
Mortality in trauma centres compared to non-trauma centres.

Ahmed 2019 reports an adjusted odds ratio (AOR) of 0.91 (95% CI 0.80 - 1.04) for higher-level care compared to lower-level care, concluding that higher-level care failed to show any mortality benefit.

Cook 2012 reports an AOR of 0.71 (95% CI 0.56-0.91) for level 1 care, AOR 1.17 (95% CI 0.90 - 1.51) for level 2 care and AOR 1.22 (95% CI 0.90 - 1.66) level 3 and 4 care. The authors conclude that there was a lower adjusted odds ratio of mortality in level 1 centers compared to level 2 and level 3 or 4 centers.

Roubik 2017 reports an observed: expected mortality of 1.001 (95% CI 1.001 - 1.002) for higher-level care, and 0.973 (95% CI 0.971 - 0.975) for lower-level care. The authors conclude that, whilst their large study size may generate statistically but not clinically significant findings, the differential O:E mortality observed for level 3 and 4 centers indicates factors beyond case mix may account for this difference.

### Temporal Trends in Mortality

Roubik 2017 reports a decrease in the AOR for mortality in higher-level centers from 2008 (AOR 0.9, 95% CI 0.9 - 1.0) to 2014 (AOR 0.7, 95% CI 0.6-0.7), using 2007 as the reference year. There was no significant temporal trend in mortality at any other trauma center level.

Cook 2012 found that the 95% confidence intervals for the AORs all crossed 1, except in the year 2001. They conclude that the adjusted odds ratio for mortality over time remained consistent.

### Secondary Outcomes

Ahmed 2019 is the only study to compare secondary outcomes. The median time to patient expiration was 6 days (IQR 3-11 days, p = 0.17) and the median time to patient discharge was 5 days (IQR 4-7 days, p < 0.001) in both higher-level and lower-level care. This is comparable to the length of stay observed in Cook 2012 and Roubik 2017.

Ahmed 2019 demonstrates a statistically significant difference in the discharge disposition. In higher-level care 21.1% of patients (n = 3,785) were discharged home without help compared to 18.3% (n = 3290) of patients in lower-level care (p<0.001).

## Discussion

### Summary of Results

This is the first study to systematically review the effect of higher-level trauma center care on outcomes in patients who have sustained low-energy trauma. The results were divergent, with the studies reporting either no difference in outcome; an advantage in higher-level care; and an advantage in lower-level care. All studies were observational and at moderate-to-severe risk of bias so therefore these results should be interpreted with caution.

### Interpretation of Findings

Roubik 2017 is the largest study in this review and was judged to be at the least risk of bias. The finding of increased O:E mortality in level 1 and 2 trauma centers is statistically but not clinically significant. Their observation of a 2.7% (2.5% - 2.9%) diminished O:E mortality in level 3 and 4 trauma centers may be clinically significant. However, since the authors also demonstrated superior data quality at higher-level centers, which contributed 90.6% of their patient cohort, this observation may reflect less accurate performance of their model in lower-level centers.

The cohort of patients in Ahmed 2019 comprises an over-65 years subset of the Roubik 2017 study. The finding of no difference in mortality between higher- and lower-level care (AOR 0.91, 95% CI 0.80-1.04) may suggest that this relatively homogenous group of patients had no potential to benefit from the enhanced services provided by higher-level trauma care.

Cook 2012 is the smallest patient cohort in this review and pertains to only a small number of hospitals in one geographical area. The finding of reduced mortality (AOR 0.71, 95% CI 0.56 - 0.91) pertains only to a single level 1 trauma center (compared to 141 in Roubik 2017) and the outcomes of 803 patients. It is therefore difficult to draw generalizable conclusions from this result.

### Relationship of Findings to Previous Research

The lack of consistent results in these studies and the failure to demonstrate improved mortality among patients in optimally equipped centers warrants further attention.

Firstly, it is possible that there is truly no mortality benefit in higher-level care. This could be explained by the summation of patient- and injury-related factors resulting in a disease process which even optimally resourced facilities are unable to ameliorate. The best support for this hypothesis is provided by Ahmed 2019, who found no mortality benefit in a cohort closely representing the emerging population of low-energy trauma patients in terms of age, mechanism of injury and absence of physiological derangement. ^27^ However, Ahmed 2019’s finding that 2.8% more patients were discharged home from higher-level centers implies the contrary: that this patient population may benefit from higher-level care, but those benefits are realized in functional outcomes rather than mortality.

Secondly, it is possible that current comparisons of injury severity are less accurate in the low-energy trauma population. For example, one flaw of the Injury Severity Score is that multiple injuries in one body region are under-represented in the overall score.^33^ This may have a disproportionate impact in the low-energy trauma population, who suffer a high proportion of head injuries, potentially resulting in under-recognition of their injury burden.^13^ An alternative definition of major trauma has been proposed based on the receipt of life-saving interventions and consequently the need for higher-level care.^34,35^

Thirdly, the timing or nature of outcome measurement in this patient population may be sub-optimal. The standard outcome of 30-day mortality may fail to measure trauma-related deaths in older patients: although most deaths among severely injured older trauma patients (ISS >15, age ≥75) occur within 20 days,^36^ it is possible that many deaths in less severely-injured patients occur later through the accumulation of complications. In addition, trauma frequently entails morbidity and impaired quality of life among survivors.^37^ Patient-reported outcome measures, such as those now collected by TARN, may be at least as appropriate in assessing the efficacy of trauma centers as current standards.

Finally, the findings of this study are supportive of a more general hypothesis: that patients injured by low-energy trauma have distinct care needs, and that specialization of trauma care may not be the most appropriate solution in low-energy trauma. This hypothesis is supported by several studies which demonstrate improved outcomes when extra resources are dedicated to prompt, comprehensive assessment and holistic management of low-energy trauma patients.^16,17,38,39^

### Implications for Future Research

This result demonstrates that the role of higher-level trauma centers in the care of patients suffering low-energy trauma requires further research. Studies at lower risk of bias and studies in non-US trauma systems and populations are required. In addition, further research is also needed into the potential of specific interventions to improve outcomes in this population.

Other areas of importance include the management of low-energy trauma patients with (1) specific injury patterns such as intracranial or thoracic injuries; (2) patients in different injury severity categories, (3) patients who require transfer between lower- and higher-level care, and (4) the examination of outcomes other than in-patient mortality, for example patient quality of life. In relation to the latter, there is also a lack of qualitative research in this field exploring the values and attitudes of older trauma patients to receiving care in potentially very distant hospitals compared to their local hospital.

### Strengths and Limitations

Comparing trauma centers using their ACS verification level has face validity owing to the “homogeneity of resources and expectations of clinical practice”.^20,29^ However, it has been demonstrated that significant differences in risk-adjusted mortality outcomes exist between similarly verified ACS trauma centres.^40^. The potential impact of intra-level variation in care is mitigated in this review by the large numbers of hospitals in two of the included studies.

A closely related issue is that the generalizability of the findings of this review to non-ACS-verified trauma centers and to hospitals outside the USA is limited. International variation in other aspects of healthcare provision, notably pre-hospital medical services, as well as differences in population demographics and injury patterns are also likely to limit generalizability.

The precision of this review is reduced by the exclusion of studies of patients with specific injury patterns or grades of injury severity. For example, significant bodies of evidence exist concerning the optimum management of patients with traumatic brain injuries or a high total injury burden.^41–43^

Articles not published in English were excluded from this review which may have introduced bias against evidence from trauma systems in some high-income countries, notably in Europe and South-East Asia. Language bias against lower-to-middle-income countries is likely to be minimal because studies in these settings were explicitly excluded in the review protocol.

Finally, no formal assessment for publication bias was undertaken.

## Conclusion

Three observational studies at moderate-to-severe risk of bias found no consistent association between higher-level trauma center care and improved outcomes compared to lower-level trauma center care in adults admitted to hospital after ground-level falls. Further research at lower risk of bias and in settings outside the USA is required.

## Data Availability

The authors confirm that the data supporting the findings of this study are available within the article and its supplementary materials.

## Ethical approval

Ethical approval was not required for this systematic review.

## Guarantor

Professor Lecky is guarantor of this work.

## Contributorship

MT and FL conceived and designed this work. MT, LS, DB and AW performed data collection. All authors contributed to data analysis and interpretation. MT drafted the article. All authors contributed to critical revision and final approval of this manuscript.

## Acknowledgements

The authors would like to acknowledge the assistance provided by Anthea Tucker, health sciences librarian at the University of Sheffield, for her role in refining the search strategy.

