## Supplemental Table 1 for "Is higher-level trauma center care associated with better outcomes in patients injured by low-energy trauma?"

### Appendix 1: Search Strategies

| **Table S1.1: Medline via Ovid** | | |
| --- | --- | --- |
| **Search** | **Terms** | **Rationale** |
| 1 | Trauma Centers/ | Intervention: MTCs |
| 2 | "trauma cent*".mp |  |
| 3 | "low energy trauma".mp | Low Energy Trauma (LET) |
| 4 | Accidental falls/ | Falls |
| 5 | ("fall*" or "low fall*" or "ground level fall*" or "same level fall*" or "low velocity fall*" or "low impact fall*" or "standing height").mp |  |
| 6 | (1 or 2) and (3 or 4 or 5) | Unfiltered, unlimited results |
| 7 | Child/ or Adolescent/ or Infant/ | Paediatric filter |
| 8 | (child* or adolescen* or infan*).mp. |  |
| 9 | 7 or 8 |  |
| 10 | Developing countries.ti,ab,kf,sh | LMIC filter |
| 11 | (Africa or Asia or Caribbean or West Indies or South America or Latin America or Central America).ti,ab,kf,hw,cp |  |
| 12 | (Afghanistan or Albania or Algeria or Angola or Antigua or Barbuda or Argentina or Armenia or Armenian or Aruba or Azerbaijan or Bahrain or Bangladesh or Barbados or Benin or Byelarus or Byelorussian or Belarus or Belorussian or Belorussia or Belize or Bhutan or Bolivia or Bosnia or Herzegovina or Hercegovina or Botswana or Brasil or Brazil or Bulgaria or Burkina Faso or Burkina Fasso or Upper Volta or Burundi or Urundi or Cambodia or Khmer Republic or Kampuchea or Cameroon or Cameroons or Cameron or Camerons or Cape Verde or Central African Republic or Chad or Chile or China or Colombia or Comoros or Comoro Islands or Comores or Mayotte or Congo or Zaire or Costa Rica or Cote d'Ivoire or Ivory Coast or Croatia or Cuba or Cyprus or Czechoslovakia or Czech Republic or Slovakia or Slovak Republic or Djibouti or French Somaliland or Dominica or Dominican Republic or East Timor or East Timur or Timor Leste or Ecuador or Egypt or United Arab Republic or El Salvador or Eritrea or Estonia or Ethiopia or Fiji or Gabon or Gabonese Republic or Gambia or Gaza or Georgia Republic or Georgian Republic or Ghana or Gold Coast or Greece or Grenada or Guatemala or Guinea or Guam or Guiana or Guyana or Haiti or Honduras or Hungary or India or Maldives or Indonesia or Iran or Iraq or Isle of Man or Jamaica or Jordan or Kazakhstan or Kazakh or Kenya or Kiribati or Korea or Kosovo or Kyrgyzstan or Kirghizia or Kyrgyz Republic or Kirghiz or Kirgizstan or Lao PDR or Laos or Latvia or Lebanon or Lesotho or Basutoland or Liberia or Libya or Lithuania or Macedonia or Madagascar or Malagasy Republic or Malaysia or Malaya or Malay or Sabah or Sarawak or Malawi or Nyasaland or Mali or Malta or Marshall Islands or Mauritania or Mauritius or Agalega Islands or Mexico or Micronesia or Middle East or Moldova or Moldovia or Moldovian or Mongolia or Montenegro or Morocco or Ifni or Mozambique or Myanmar or Myanma or Burma or Namibia or Nepal or Netherlands Antilles or New Caledonia or Nicaragua or Niger or Nigeria or Northern Mariana Islands or Oman or Muscat or Pakistan or Palau or Palestine or Panama or Paraguay or Peru or Philippines or Philipines or Phillipines or Phillippines or Poland or Portugal or Puerto Rico or Romania or Rumania or Roumania or Russia or Russian or Rwanda or Ruanda or Saint Kitts or St Kitts or Nevis or Saint Lucia or St Lucia or Saint Vincent or St Vincent or Grenadines or Samoa or Samoan Islands or Navigator Island or Navigator Islands or Sao Tome or Saudi Arabia or Senegal or Serbia or Montenegro or Seychelles or Sierra Leone or Slovenia or Sri Lanka or Ceylon or Solomon Islands or Somalia or South Africa or Sudan or Suriname or Surinam or Swaziland or Syria or Tajikistan or Tadzhikistan or Tadjikistan or Tadzhik or Tanzania or Thailand or Togo or Togolese Republic or Tonga or Trinidad or Tobago or Tunisia or Turkey or Turkmenistan or Turkmen or Uganda or Ukraine or Uruguay or USSR or Soviet Union or Union of Soviet Socialist Republics or Uzbekistan or Uzbek or Vanuatu or New Hebrides or Venezuela or Vietnam or Viet Nam or West Bank or Yemen or Yugoslavia or Zambia or Zimbabwe or Rhodesia).ti,ab,kf,hw,cp |  |
| 13 | 10 or 11 or 12 |  |
| 14 | 6 not (9 or 13) | Filters applied |
| 15 | limit 14 to (english language and humans and yr="1987 -Current") | Limits applied |

| **Table S1.2: Embase via Ovid** | | |
| --- | --- | --- |
| **Search** | **Terms** | **Rationale** |
| 1 | emergency health service/ | Intervention: MTCs |
| 2 | "trauma cent*".mp |  |
| 3 | "low energy trauma".mp | Low Energy Trauma (LET) |
| 4 | falling/ | Falls |
| 5 | ("fall*" or "low fall*" or "ground level fall*" or "same level fall*" or "low velocity fall*" or "low impact fall*" or "standing height").mp |  |
| 6 | (1 or 2) and (3 or 4 or 5) | Unfiltered, unlimited results |
| 7 | child/ or adolescent/ or infant/ | Paediatric filter |
| 8 | (child* or adolesc* or infan*).mp |  |
| 9 | 7 or 8 |  |
| 10 | Developing countries.ti,ab,kw,sh | LMIC filter |
| 11 | (Africa or Asia or Caribbean or West Indies or South America or Latin America or Central America).ti,ab,kw,sh,cp |  |
| 12 | (Afghanistan or Albania or Algeria or Angola or Antigua or Barbuda or Argentina or Armenia or Armenian or Aruba or Azerbaijan or Bahrain or Bangladesh or Barbados or Benin or Byelarus or Byelorussian or Belarus or Belorussian or Belorussia or Belize or Bhutan or Bolivia or Bosnia or Herzegovina or Hercegovina or Botswana or Brasil or Brazil or Bulgaria or Burkina Faso or Burkina Fasso or Upper Volta or Burundi or Urundi or Cambodia or Khmer Republic or Kampuchea or Cameroon or Cameroons or Cameron or Camerons or Cape Verde or Central African Republic or Chad or Chile or China or Colombia or Comoros or Comoro Islands or Comores or Mayotte or Congo or Zaire or Costa Rica or Cote d'Ivoire or Ivory Coast or Croatia or Cuba or Cyprus or Czechoslovakia or Czech Republic or Slovakia or Slovak Republic or Djibouti or French Somaliland or Dominica or Dominican Republic or East Timor or East Timur or Timor Leste or Ecuador or Egypt or United Arab Republic or El Salvador or Eritrea or Estonia or Ethiopia or Fiji or Gabon or Gabonese Republic or Gambia or Gaza or Georgia Republic or Georgian Republic or Ghana or Gold Coast or Greece or Grenada or Guatemala or Guinea or Guam or Guiana or Guyana or Haiti or Honduras or Hungary or India or Maldives or Indonesia or Iran or Iraq or Isle of Man or Jamaica or Jordan or Kazakhstan or Kazakh or Kenya or Kiribati or Korea or Kosovo or Kyrgyzstan or Kirghizia or Kyrgyz Republic or Kirghiz or Kirgizstan or Lao PDR or Laos or Latvia or Lebanon or Lesotho or Basutoland or Liberia or Libya or Lithuania or Macedonia or Madagascar or Malagasy Republic or Malaysia or Malaya or Malay or Sabah or Sarawak or Malawi or Nyasaland or Mali or Malta or Marshall Islands or Mauritania or Mauritius or Agalega Islands or Mexico or Micronesia or Middle East or Moldova or Moldovia or Moldovian or Mongolia or Montenegro or Morocco or Ifni or Mozambique or Myanmar or Myanma or Burma or Namibia or Nepal or Netherlands Antilles or New Caledonia or Nicaragua or Niger or Nigeria or Northern Mariana Islands or Oman or Muscat or Pakistan or Palau or Palestine or Panama or Paraguay or Peru or Philippines or Philipines or Phillipines or Phillippines or Poland or Portugal or Puerto Rico or Romania or Rumania or Roumania or Russia or Russian or Rwanda or Ruanda or Saint Kitts or St Kitts or Nevis or Saint Lucia or St Lucia or Saint Vincent or St Vincent or Grenadines or Samoa or Samoan Islands or Navigator Island or Navigator Islands or Sao Tome or Saudi Arabia or Senegal or Serbia or Montenegro or Seychelles or Sierra Leone or Slovenia or Sri Lanka or Ceylon or Solomon Islands or Somalia or South Africa or Sudan or Suriname or Surinam or Swaziland or Syria or Tajikistan or Tadzhikistan or Tadjikistan or Tadzhik or Tanzania or Thailand or Togo or Togolese Republic or Tonga or Trinidad or Tobago or Tunisia or Turkey or Turkmenistan or Turkmen or Uganda or Ukraine or Uruguay or USSR or Soviet Union or Union of Soviet Socialist Republics or Uzbekistan or Uzbek or Vanuatu or New Hebrides or Venezuela or Vietnam or Viet Nam or West Bank or Yemen or Yugoslavia or Zambia or Zimbabwe or Rhodesia).ti,ab,kw,sh,cp |  |
| 13 | 10 or 11 or 12 |  |
| 14 | 6 not (9 or 13) | Filters applied |
| 15 | limit 14 to (human and english language and yr="1987 -Current") | Limits applied |

| **Table S1.3: CINHAL via EBSCO** | | |
| --- | --- | --- |
| **Search** | **Terms** | **Rationale** |
| 1 | MH Trauma Centers | Intervention: MTCs |
| 2 | TI or AB or SU "trauma cent*" |  |
| 3 | Ti or AB or SU "low energy trauma" | Low Energy Trauma (LET) |
| 4 | MH Accidental falls | Falls |
| 5 | TI or AB or SU ("fall*" or "low fall*" or "ground level fall*" or "same level fall*" or "low velocity fall*" or "low impact fall*" or "standing height") |  |
| 6 | S1 or S2 | Unfiltered, unlimited results |
| 7 | S3 or S4 or S5 |  |
| 8 | S6 and S7 |  |
| 9 | MH (Child or Adolescence or Infant) | Paediatric filter |
| 10 | TI or AB or SU (child* or adolescen* or infan*) |  |
| 11 | 9 or 10 |  |
| 12 | MH or TI or AB or SU "Developing countries" | LMIC filter |
| 13 | MW or TI or AB or SU (Africa or Asia or Caribbean or West Indies or South America or Latin America or Central America) |  |
| 14 | (Afghanistan or Albania or Algeria or Angola or Antigua or Barbuda or Argentina or Armenia or Armenian or Aruba or Azerbaijan or Bahrain or Bangladesh or Barbados or Benin or Byelarus or Byelorussian or Belarus or Belorussian or Belorussia or Belize or Bhutan or Bolivia or Bosnia or Herzegovina or Hercegovina or Botswana or Brasil or Brazil or Bulgaria or Burkina Faso or Burkina Fasso or Upper Volta or Burundi or Urundi or Cambodia or Khmer Republic or Kampuchea or Cameroon or Cameroons or Cameron or Camerons or Cape Verde or Central African Republic or Chad or Chile or China or Colombia or Comoros or Comoro Islands or Comores or Mayotte or Congo or Zaire or Costa Rica or Cote d'Ivoire or Ivory Coast or Croatia or Cuba or Cyprus or Czechoslovakia or Czech Republic or Slovakia or Slovak Republic or Djibouti or French Somaliland or Dominica or Dominican Republic or East Timor or East Timur or Timor Leste or Ecuador or Egypt or United Arab Republic or El Salvador or Eritrea or Estonia or Ethiopia or Fiji or Gabon or Gabonese Republic or Gambia or Gaza or Georgia Republic or Georgian Republic or Ghana or Gold Coast or Greece or Grenada or Guatemala or Guinea or Guam or Guiana or Guyana or Haiti or Honduras or Hungary or India or Maldives or Indonesia or Iran or Iraq or Isle of Man or Jamaica or Jordan or Kazakhstan or Kazakh or Kenya or Kiribati or Korea or Kosovo or Kyrgyzstan or Kirghizia or Kyrgyz Republic or Kirghiz or Kirgizstan or Lao PDR or Laos or Latvia or Lebanon or Lesotho or Basutoland or Liberia or Libya or Lithuania or Macedonia or Madagascar or Malagasy Republic or Malaysia or Malaya or Malay or Sabah or Sarawak or Malawi or Nyasaland or Mali or Malta or Marshall Islands or Mauritania or Mauritius or Agalega Islands or Mexico or Micronesia or Middle East or Moldova or Moldovia or Moldovian or Mongolia or Montenegro or Morocco or Ifni or Mozambique or Myanmar or Myanma or Burma or Namibia or Nepal or Netherlands Antilles or New Caledonia or Nicaragua or Niger or Nigeria or Northern Mariana Islands or Oman or Muscat or Pakistan or Palau or Palestine or Panama or Paraguay or Peru or Philippines or Philipines or Phillipines or Phillippines or Poland or Portugal or Puerto Rico or Romania or Rumania or Roumania or Russia or Russian or Rwanda or Ruanda or Saint Kitts or St Kitts or Nevis or Saint Lucia or St Lucia or Saint Vincent or St Vincent or Grenadines or Samoa or Samoan Islands or Navigator Island or Navigator Islands or Sao Tome or Saudi Arabia or Senegal or Serbia or Montenegro or Seychelles or Sierra Leone or Slovenia or Sri Lanka or MW or TI or AB or SU (Ceylon or Solomon Islands or Somalia or South Africa or Sudan or Suriname or Surinam or Swaziland or Syria or Tajikistan or Tadzhikistan or Tadjikistan or Tadzhik or Tanzania or Thailand or Togo or Togolese Republic or Tonga or Trinidad or Tobago or Tunisia or Turkey or Turkmenistan or Turkmen or Uganda or Ukraine or Uruguay or USSR or Soviet Union or Union of Soviet Socialist Republics or Uzbekistan or Uzbek or Vanuatu or New Hebrides or Venezuela or Vietnam or Viet Nam or West Bank or Yemen or Yugoslavia or Zambia or Zimbabwe or Rhodesia). |  |
| 15 | S12 or S13 or S14 |  |
| 16 | S11 or S15 | Filters combined |
| 17 | 8 not 16 AND Limits (english language and humans and yr="1987 -Current") | Filters and limits applied |

| **Table S1.4: Cochrane Library** | | |
| --- | --- | --- |
| **Search** | **Terms** | **Rationale** |
| 1 | MeSH descriptor: [Trauma Centers] explode all trees | Trauma centers |
| 2 | "trauma cent*" |  |
| 3 | "low energy trauma" | Low energy trauma |
| 4 | MeSH descriptor: [Accidental Falls] explode all trees | Falls |
| 5 | ("fall*" or "low fall*" or "ground level fall*" or "same level fall*" or "low velocity fall*" or "low impact fall*" or "standing height") |  |
| 6 | (#1 or #2) and (#3 or #4 or #5) | Unfiltered results |

| **Table S1.5 Open Grey** | | |
| --- | --- | --- |
| **Search** | **Terms** | **Rationale** |
| 1 | Trauma center discipline:(06E - Medicine) | Limited search facility |

| **Table S1.6: Grey Literature Report** | | |
| --- | --- | --- |
| **Search** | **Terms** | **Rationale** |
| 1 | trauma center | Limited search facility |

### Appendix 2: Studies Excluded at Full Paper Stage

| **Table S2.1: Studies excluded at global quality assessment** | | | | |
| --- | --- | --- | --- | --- |
| Study | Does this study compare outcomes at a major trauma centre and a non-trauma centre? | Does this study report outcomes in patients with low-energy trauma? | Decision | Justification |
| Ahmed  2019b  (28) | Yes | Yes | Exclude | This study cohort comprises all the 40,800 patients in Ahmed 2019 (30). Whilst outcomes at different levels of trauma centre are reported, the aim of the paper is to produce a risk-stratification model. The findings are better described in the included paper. |
| Ahmed  2019c  (29) | Yes | Yes | Exclude | This study is a cohort of 15,256 patients with thoracic injuries drawn from the same cohort as Ahmed 2019 (30). After propensity matching the authors found no mortality difference between patients treated at ACS-verified level 1 & 2 cantres compared to ACS-verified level 3 & 4 centres (mortality 4.4% vs 3.9%, p = 0.14). |
| Scheetz  2015  (31) | Yes | Yes | Exclude | This retrospective cohort identified 3,331 patients with traumatic brain injury following same-level falls using a New York State healthcare database. Among other goals, it aimed to identify predictors of short term mortality. The presented logistic regression model of mortality reported an OR for mortality of 1.49 for trauma centre admission (95% CI 1.17-1.89).  The review was excluded for the following reasons:  - Patients with missing trauma centre status were removed from the analysis.The number removed is not reported.  - Interfacility transfer patients were counted only upon discharge from the initial hospital, introducing significant risk of tertiary referral bias.  - The variables in the logistic regression model in text and table are discrepant.  - The referent standard used for comparison of traumatic brain injury severity was a composite of ‘unclassified’ injuries. |

### Appendix 3: Multiple Logistic Regression Models

| **Table S4.1: Multiple regression model from Ahmed 2019** | | | |
| --- | --- | --- | --- |
| Calibration and discrimination | NR* | | |
| Variable | Odds Ratio | 95% CI | P |
| Age | NR* | NR* | NR* |
| Sex |  |  |  |
| Race/ethnicity |  |  |  |
| Systolic blood pressure |  |  |  |
| Heart rate |  |  |  |
| Respiratory rate |  |  |  |
| Injury severity score |  |  |  |
| *Author contacted but information not available | | | |

| **Table S4.2: Multiple regression model from Cook 2012** | | | |
| --- | --- | --- | --- |
| Calibration and discrimination | AUROC 0.85, Hosmer-Lemeshow chi-square 4.98, p = 0.89 | | |
| Variable | Odds Ratio | 95% CI | P |
| Hypotension | 8.91 | 6.09-13.05 | <0.001 |
| Metabolic acidosis | 7.19 | 2.36-21.88 | 0.001 |
| Comorbid cancer | 3.83 | 1.57-9.35 | 0.003 |
| Comorbid renal disorder | 3.25 | 1.30-8.13 | 0.01 |
| Comorbid cardiac valve disorder | 2.53 | 1.04_6.15 | 0.04 |
| Comorbid congestive heart failure | 2.45 | 1.58-3.79 | <0.001 |
| Comorbid cardiac arrhythmia | 2.24 | 1.32-3.80 | 0.003 |
| Isolated traumatic brain injury | 2.24 | 1.13_4.43 | 0.02 |
| Any traumatic brain injury | 1.95 | 1.43-2.66 | <0.001 |
| Male gender | 1.74 | 1.17-2.58 | 0.006 |
| Trauma Mortality Prediction Model score | 1.58 | 1.27-1.95 | <0.001 |
| Age | 1 .04 | 1.02-1.05 | <0.001 |
| Glasgow Coma Scale score | 0.76 | 0.71-0.82 | <0.001 |

| **Table S4.3: Multiple regression model from Roubik 2017** | | | |
| --- | --- | --- | --- |
| Calibration and discrimination | AUROC 0.90 | | |
| Variable | Odds Ratio | 95% CI | P |
| Do not Resuscitate (DNR) status | 3.82 | (3.66-3.98) | <0.001 |
| Disseminated cancer | 2.9 | (2.71-3.11) |  |
| Ascites within 30 days/cirrhosis | 2.89 | (2.59-3.23) |  |
| Chemotherapy for cancer | 2.11 | (1.72-2.57) |  |
| Currently requiring dialysis | 1.79 | (1.67-1.92) |  |
| Congestive heart failure | 1.63 | (1.58-1.69) |  |
| Chronic respiratory disease | 1.4 | (1.33-1.46) |  |
| Bleeding disorder | 1.24 | (1.20-1.28) |  |
| Hypertension requiring medication | 0.86 | (0.84-0.88) |  |
| Cervical spine fracture | 1.43 | (1.36-1.49) |  |
| Thoracic spine fracture | 1.4 | (1.31-1.50) |  |
| Sacral/coccyx fracture | 1.33 | (1.18-1.49) |  |
| Lumbar spine fracture | 1.3 | (1.20-1.41) |  |
| Lower extremity fracture | 1.21 | (1.15-1.28) |  |
| Shoulder/upper arm injury | 1.17 | (1.12-1.23) |  |
| Hip injury | 1.17 | (1.11-1.23) |  |
| Brain injury | 1.11 | (1.06-1.15) |  |
| Chest injury | 1.08 | (1.03-1.13) | 0.002 |
| Facial fracture | 0.9 | (0.86-0.95) | <0.001 |
| Skull fracture | 0.89 | (0.84-0.94) |  |
| ICU admission | 4.23 | (4.10-4.36) |  |
| Glasgow Coma Scale <15 | 2.92 | (2.84-3.00) |  |
| Systolic blood pressure (mmHg) | 1.83 | (1.69-1.98) |  |
| Transfer from outside hospital | 0.82 | (0.80-0.85) |  |
| Alcohol on board on amival | 0.71 | (0.66-0.76) |  |
| Cardiac arrest with cardiopulmonary resuscitation | 25.74 | (23.54-28.15) |  |
| Stroke | 3.62 | (3.22-4.08) |  |
| Severe sepsis | 3.16 | (2.87-3.48) |  |
| Acute lung injury/acute respiratory distress syndrome | 3.1 | (2.89-3.33) |  |
| Acute kidney injury | 2.66 | (2.49-2.85) |  |
| Myocardial infarction | 2.43 | (2.22-2.66) |  |
| Unplanned intubation | 2.32 | (2.13-2.52) |  |
| Pulmonary embolism | 2.13 | (1.86-2.44) |  |
| Pneumonia | 1.88 | (1.78-1.98) |  |
| Drug/alcohol withdrawal | 0.44 | (0.37-0.51) |  |
| 2008 | 0.94 | (0.87-1.01) | 0.09 |
| 2009 | 0.82 | (0.77-0.88) | <0.001 |
| 2010 | 0.81 | (0.75-0.86) |  |
| 2011 | 0.73 | (0.68-0.78) |  |
| 2012 | 0.69 | (0.64-0.74) |  |
| 2013 | 0.65 | (0.61-0.69) |  |
| 2014 | 0.67 | (0.63-0.72) |  |

### Appendix 4: Tables Including State Trauma Centre

| Table 4: Mortality in trauma centres compared to non-trauma centres | | | |
| --- | --- | --- | --- |
| Study | Trauma centre | Unadjusted Mortality  n (%) | Adjusted Comparison for Mortality  Summary statistic (95% CI) |
| Ahmed  2019  (30) | ACS 1 & 2 | NR* | AOR 0.91 (0.80-1.04) |
|  | ACS 3 & 4 |  | Reference standard |
| Cook  2012  (32) | ACS 1 | 48 (6%) | AOR 0.71 (0.56-0.91) |
|  | ACS 2 | 69 (3.1%) | AOR 1.17 (0.90 - 1.51) |
|  | ACS 3 & 4 | 59 (1.1%) | AOR 1.22 (0.90 - 1.66) |
| Roubik  2017  (33) | ACS 1 & 2 | 22,382 (4.45%) | O:E 1.001 (1.001-1.002) |
|  | State 1 & 2 | 9,122 (4.3%) | O:E 0.998 (0.998-0.999) |
|  | ACS 3 & 4 | 1,342 (2.54%) | O:E 0.973 (0.971-0.975) |
|  | State 3 & 4 | 1,418 (3.11%) | O:E 1.043 (1.041-1.044) |
| *Author contacted but information not available | | | |
